# A Real-Time Deep Learning Approach for Inferring Intracranial Pressure from Routinely Measured Extracranial Waveforms in the Intensive Care Unit

**DOI:** 10.1101/2023.05.16.23289747

**Authors:** Shiker S. Nair, Alina Guo, Joseph Boen, Ataes Aggarwal, Ojas Chahal, Arushi Tandon, Meer Patel, Sreenidhi Sankararaman, Tej Azad, Romain Pirracchio, Robert D. Stevens

**Affiliations:** Department of Biomedical Engineering, Johns Hopkins University Whiting School of Engineering, Baltimore, MD, USA; Department of Neurosurgery, Johns Hopkins University School of Medicine, Baltimore, MD, USA; Department of Anaesthesia and Perioperative Care, UCSF, San Francisco, CA, USA; Departments of Anesthesiology and Critical Care Medicine, Neurology, and Radiology, Johns Hopkins School of Medicine, Baltimore, MD, USA

**Keywords:** Arterial blood pressure, deep learning, electrocardiogram, intracranial pressure, noninvasive diagnosis, photoplethysmography, precision medicine

## Abstract

**Objective:** Intracranial pressure (ICP) is a physiological variable used to assess the neurological state of patients with life-threatening intracranial pathology, such as traumatic brain injury or stroke. The current standard of care for measuring ICP requires a catheter to be inserted into the brain, which is associated with an appreciable risk of hemorrhage and infection. We hypothesize that ICP can be computed from extracranial waveforms routinely measured in the Intensive Care Unit (ICU), such as invasive arterial blood pressure (ABP), photoplethysmography (PPG), and electrocardiography (ECG).

**Methods:** We extracted 600 hours of simultaneous ABP, ECG, PPG, and ICP data (sampled at 125 Hz) across 10 different patients from the MIMIC III Waveform Database. These recordings were segmented into 10 second windows and used to train six different deep learning models with ABP, ECG, and PPG waveforms as input features. Models were evaluated in both a single-patient analysis and multi-patient analysis.

**Results:** The performances of the six deep learning models were compared, revealing two tiers of performance. Among the top-tier models, the mean average error (MAE) for inferring ICP was approximately 1.50 mmHg for single-patient analysis and 5 mmHg for multi-patient analysis.

**Conclusions:** These preliminary and novel results indicate the feasibility and accuracy of noninvasive ICP estimation by training deep learning models with extracranial physiological data. With further validation, this approach could be implemented in a continuous real-time fashion, thereby reducing risks associated with invasive monitoring and allowing more timely treatment of patients with critical brain injuries.

## I. Introduction

Traumatic brain injury (TBI), a type of brain damage caused by physical trauma to the head, is one of the leading causes of death and disability worldwide [1]. In the US alone, recent data indicate 223,135 TBI-related hospitalizations and close to 70,000 TBI-related deaths [2]. The leading cause of TBI-related death is increased intracranial pressure (ICP) [3]. ICP is defined as the pressure exerted within the cranial compartment of a patient [4]. At rest, a normal supine adult’s ICP is 0-20 mmHg [5,6]. Intracranial hypertension is defined as a sustained increase of ICP above 20 mmHg [4]. This is typically caused by an expanding intracranial mass, cerebral edema, or a process impeding cerebrospinal fluid flow and/or reabsorption through the ventricular and subarachnoid spaces [7].

ICP monitoring is highly recommended in patients with severe TBI [3]. In cases of elevated ICP, immediate treatment is required for the patient (i.e., administration of hypersaline agents) [4]. Currently, the standard for monitoring ICP is the placement of an external ventricular drain (EVD) or intraparenchymal brain monitor (IPM) [5,8]. Insertion of such devices requires a hole to be drilled in the skull and the probe to be inserted through the tissues of the brain. The invasive nature of this procedure poses risks including infection and hemorrhage. The risk of infection for the EVD is 5-14% and the risk of hemorrhage is 5-7% [3,6]. Additionally, insertion of an EVD requires a neurosurgery team and takes approximately 30-60 minutes, resulting in delayed ICP measurements potentially delaying ICP detection and intervention [9]. This could result in rapid deterioration of a TBI patient and, eventually, lead to death [4]. Thus, there remains a need for a less invasive, continuous way of monitoring ICP that is reliable and accurate.

Previous research has indicated coupling between ICP and the cardiovascular system, suggesting that information present in extracranial physiological waveforms is correlated with intracranial events [10]-[12]. Cerebral autoregulation may be impaired as a consequence of elevated ICP, leading to pathological changes in cerebrovascular vasoreactivity, cerebral blood flow, and oxygenation. Here we explore the relationship between ICP and three physiological indices which are routinely measured/monitored in the ICU: invasive arterial blood pressure monitoring (ABP), photoplethysmography (PPG), and electrocardiography (ECG). We hypothesize that an estimated ICP measure can be computed by decoding extracranial physiological waveform data.

Estimating ICP using extracranial waveforms would allow clinicians to assess patients with intracranial abnormalities in a noninvasive, cost-effective manner. Furthermore, such a solution could be unobtrusively integrated into the current clinical workflow in the ICU.

## II. Related Works

Our research presents a novel approach to predicting ICP in patients with TBI that significantly differs from existing studies in the literature. We leverage traditional machine learning and deep learning architectures to predict ICP using high-frequency (125 Hz) ABP, PPG, and ECG waveform data in real-time. Although previous groups have attempted to develop both physics-based simulations and statistical models to infer ICP, there have been limited efforts to use data-driven models to accomplish this task [13, 14].

The primary novelty of our approach lies in the use of solely extracranial signals to predict the ICP waveform in quasi-real-time, using deep learning techniques. Similar studies in this field have successfully developed methods to predict the occurrence of hypertensive events or the progression of intracranial hypertension (IH) into a potentially life-threatening condition [15]-[18]. Other works have developed deep-learning algorithms to forecast the ICP waveform [19,20]. However, all of these studies rely on using ICP as an input and therefore do not offer the potential of an extracranial, non-invasive methodology for measuring ICP. There is indeed literature for the realtime prediction of ICP using extracranial signals [13,14]; however, these studies scarcely utilize deep learning in their formula-based methods.

By focusing on non-invasive methods for measuring ICP, our research has the potential for transforming the monitoring and treatment of TBI patients, particularly in resource-limited settings. ICP could become a routinely estimated vital sign beyond the intensive care unit (ICU). In addition, real-time waveform re-creation using deep learning can be seamlessly integrated into the current monitoring workflow to replace invasive catheters while still giving clinicians the autonomy to interpret ICP waveforms.

## III. Data

MIMIC-III (v1.4, 2016) is an intensive care database that comprises de-identified data from 46,520 patients and 58,976 admissions, recorded between June 1, 2001, and October 31, 2012, at the Beth Israel Deaconess Medical Center in Boston, USA [21]. The data includes demographics, admission, and discharge notes, International Classification of Diseases 9th revision (ICD-9) codes for diagnoses, radiological findings, survival data, vital sign recordings, medications, and procedures for each patient [21]. MIMIC-III also contains high-frequency physiological waveforms data from a subset of patients. We decided to model ICP from ABP, ECG, and PPG. By choosing these waveforms, we had three modalities of predicting ICP and assessing the contributions of each waveform in the modeling tasks.

## IV. Methodology

### A. Preprocessing, Cohort Selection, and Patient Demographics

Patients who were at least 18 years old with ECG, PPG, ABP, and ICP recordings for at least five minutes were included in this study. To accomplish this, we first assessed whether a patient had recordings of all four waveforms. Because ECG is measured at multiple different leads, we used the most prevalent ECG lead (II) in our patient population of interest. The rationale behind the common usage of ECG lead II for ICP prediction is rooted in its ability to provide a reliable depiction of the mechanical forces engendered by the cardiac system and their subsequent impact on ICP [22]. After identifying patients who have ICP, ABP, ECG Lead II, and PPG data available, we removed patients who had less than five minutes of overlapping data between these four waveforms. This was done based on the knowledge that elevated ICP lasting for five minutes or more may indicate a hypertensive event, according to clinical experts [23]. Therefore, it was important to ensure that we had sufficient data to accurately detect such an event if it occurred.

From the patients that remained, the waveform data (synchronized in time) were sliced into five-minute segments. To ensure data quality, five-minute segments that contained any not a number (NaN) values or physiologically unrealistic values (ICP < -10 mmHg or > 200 mmHg, ABP < 20 mmHg or > 300 mmHg, not NaN mean heart rate) unique to each waveform were filtered out. To address potential issues with EVD calibration, an additional periodicity filtering step was applied to the ICP signals. This filtering step removed extremely highfrequency and low-frequency waveforms that could lead to inaccuracies in the data. To ensure that intracranial hypertension events were adequately represented in the data, patients who had less than one hour of intracranial hypertension recordings were excluded. For this study, an ICP hypertensive event was defined as an ICP greater than 20 mmHg sustained for at least five minutes [23]. Although recent traumatic brain injury guidelines propose a threshold of 22 mmHg, our study includes a heterogeneous population of patients who underwent ICP monitoring for both traumatic and nontraumatic brain disorders [4]. Therefore, we opted to use a threshold of 20 mmHg. Figure 1 summarizes these pre-processing workflow for cohort selection.

**FIG. 1:**
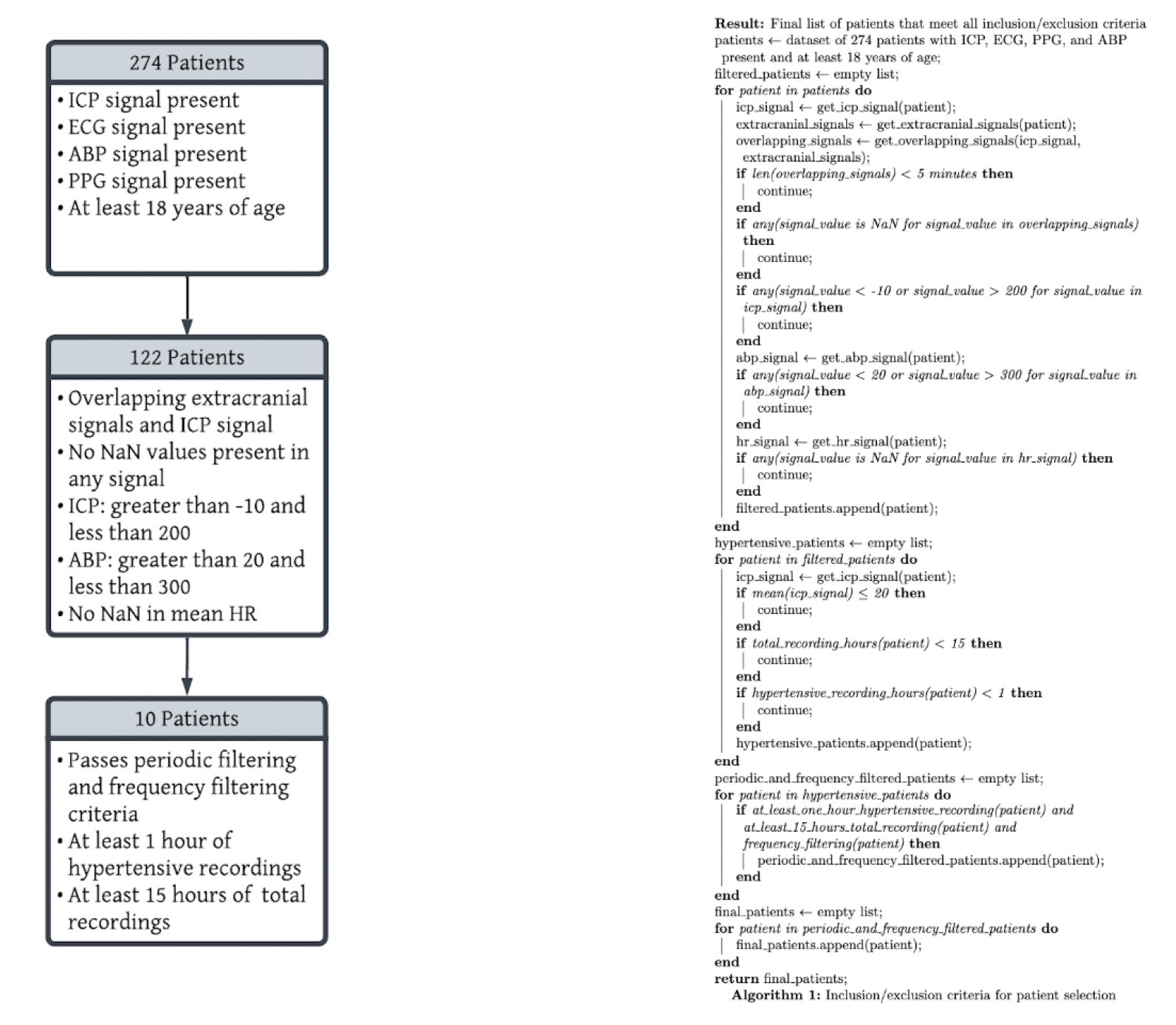
Cohort Selection Workflow (Left) and Pre-Processing Methodology (Right)

After applying this pre-processing scheme, 10 patients remained for downstream analysis containing more than 600 hours of recorded ICP, ABP, EKG, PPG data (more than 270,000,000 data points). Table I highlights key demographics features of these patients.

**Table I.** Patient Demographic Data in Study

|  |  |
| --- | --- |
| **Total Number of Subjects** | n = 10 |
| **Age Median [IQR]** | 56 [33-64] |
| **Sex(% M, % F)** | 30, 70 |
| **Admission Type (Number of Subjects)** | Emergency (8) |
|  | Urgent (2) |
| **Discharge Location (Number of Subjects)** | Rehab/Distinct Part Hosp (5) |
|  | Dead/Expired (3) |
|  | Home Health Care (1)<br>Home (1) |
| **Diagnosis (Number of Subjects)** | Subarachnoid hemorrhage (5) |
|  | Acute liver failure (2) |
|  | Traumatic brain injury (1) |
|  | Intracerebral hemorrhage (1) |
|  | Closed fracture of base of skull (1) |

### B. Determining Window Length for ICP Prediction Task

To ensure that the ICP prediction task was as close to real-time as possible, a window size of 10 seconds was chosen. To assess the amount of information lost as the window size decreased, a dynamic time warping analysis (DTW) was conducted [24]. The patient data was divided into windows of 10 seconds, 20 seconds, 30 seconds, and 60 seconds. The DTW score for each of the extracranial waveforms (ABP, ECG, and PPG) and ICP was computed between adjacent windows. This resulted in four scores (one for each waveform) for each pair of adjacent windows. The Spearman correlation between the DTW scores for each extracranial waveformand ICP was calculated (ABP/ICP, ECG/ICP, and PPG/ICP) to assess if changes in the extracranial waveforms would be reflected in ICP.

The results showed that all window sizes had a weak positive correlation between the extracranial waveforms and ICP (Table II). This justified the choice of a 10second window size to best replicate real-time analysis.

**Table II.** Dynamic Time Warping Analysis for Assessing Information Loss Across Decreasing Window Sizes

| Window Size (s) | ICP/ABP | ICP/PPG | ICP/ECG |
| --- | --- | --- | --- |
| 10 | 0.39 | 0.29 | 0.25 |
| 20 | 0.37 | 0.30 | 0.29 |
| 30 | 0.40 | 0.33 | 0.31 |
| 60 | 0.42 | 0.37 | 0.35 |

### C. Model Selection and Architecture

The goal of this study is to predict the entire ICP waveform (125 Hz) from ABP, ECG, and PPG waveforms (125 Hz) in 10-second windows. Given this is a multivariate time series prediction problem, we implemented six different deep learning models which are fit to accomplish this task and have been used in previous studies in similar problem spaces for time-series based predictions (Figure 2):

- Recurrent Neural Network (RNN) [25,26]
- Gated Recurrent Unit (GRU) [27]-[30]
- Long-Short Term Memory (LSTM) network [19], [30-32]
- Temporal Convolutional Networks (TCN) [33]-[36]
- Volume-Net (VNET) [37]
- Transformer [38,39]

**FIG. 2:**
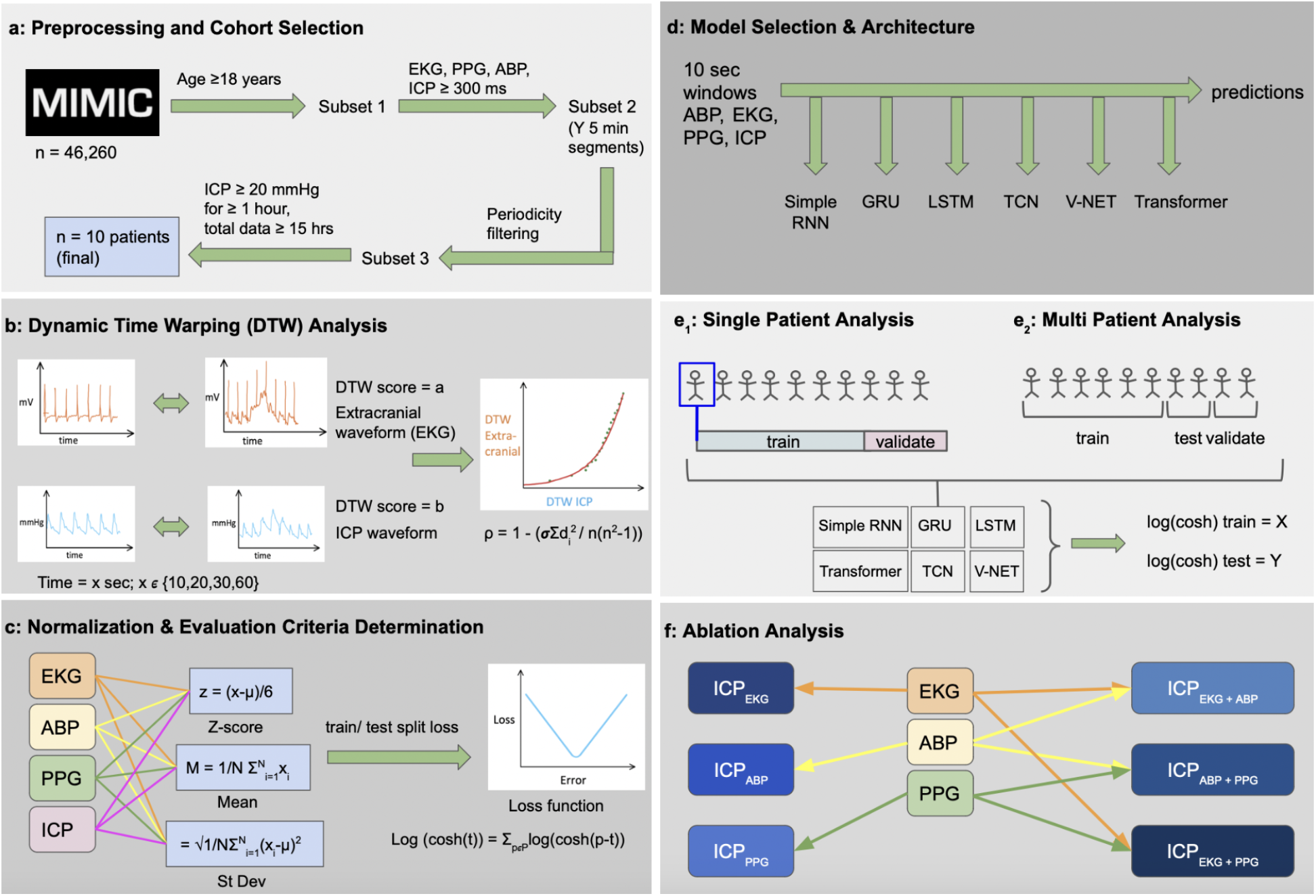
Overview of Methodology and Analysis Performed in Study

These six models vary in sophistication and complexity. In general, adding more layers improved the predictions for each model, however, this increased the risk of overfitting. Rigorous hyperparameter tuning was applied for each model, specifically finding an optimal learning rate, batch size, and number of nodes for each layer.

### D. Normalization and Evaluation Criteria

Both features (ABP, ECG, and PPG) and labels (ICP) waveforms were independently normalized for the prediction tasks. Normalization was completed using a standard scaling function (subtracting by mean and dividing by the standard deviation of the waveform) which was applied to all the data before splitting it into a training and testing set. While it is common practice to normalize the training set and apply this normalization scheme to the test set, the small number of patients used in this analysis (n=10) did not allow for a generalizable normalization function in the training set that would properly transfer over to the testing set. As a result, all the data was normalized before splitting into a training and testing set. Predictions were then unscaled back into a clinically interpretable ICP using the inverse scaling function.

Different loss functions were assessed when optimizing each model and it was determined that the log hyperbolic cosine (log cosh) loss was the best optimization function based on convergence. It is defined as the logarithm of the hyperbolic cosine of the difference between the true (t) and predicted values (p) (Equation 1). The logcosh loss function is a smooth approximation of the MAE loss function, allowing it to be easily optimized through gradient descent methods. Furthermore, logcosh is known to converge faster than other loss functions such as MAE, overall making it an ideal function for deep learning tasks [40]-[42]. MAE and root mean squared error (RMSE) were also metrics used to report model performance (Equation 2 and Equation 3).

Equation 1. Logcosh Loss Function Formula

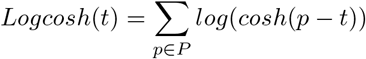

Equation 2. Mean Average Error Function Formula

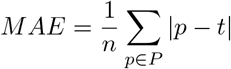

Equation 3. Root Mean Squared Error Formula

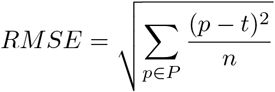

### E. Single-Patient Analysis

The first experiment conducted evaluated the performance of each of the six models within a single-patient for each of the 10 patients in the study. The goal of this experiment was to ascertain that each model was capable of predicting ICP waveforms from the extracranial waveforms in the simplest context: within a single-patient. This removes any notion of generalizability and enables the comparison of each model in a simpler context before considering inter-patient generalization. Data were split into 70% train/validation set and 30% test set. The mean and median logcosh for training and testing datasets were reported for each model along with the standard deviation. Using this model information, we selected the top model(s) to run a multi-patient analysis. Additional follow-up analysis was performed to understand the strengths and weaknesses of each model and trends in error and prediction.

### F. Multi-Patient Analysis on Top Models

To ensure that the top models are generalizable to new patients, a multi-patient analysis was conducted using 6 patients for training, 2 for validation, and 2 for the test set. This approach ascertains that the models are evaluated on data from patients that are distinct from those used in the training set, thus providing a robust assessment of model performance. The mean and median logcosh values for the training and testing datasets were reported for each model, to provide a measure of model performance accuracy and consistency. Furthermore, additional follow-up analyses were performed to understand trends across the top models and across different patients in the validation and testing sets. This analysis is crucial for ensuring that the models are generalizable and can be applied to new patients in clinical practice, where the algorithm will be trained on different patients than the patients it will be used on in the ICU.

### G. Ablation Analysis on Multi-Patient Top Model

To assess extracranial waveform importance, an ablation study was conducted in the top model for multipatient analysis. All subsets of input extracranial waveforms were tested: ABP only, PPG only, ECG only, ABP + PPG, PPG + ECG, and ABP + ECG. The goal of this study was to understand the effect that dropping extracranial waveforms had on model performance.

### H. Clinical Error Analysis Procedure

To improve the accuracy of predictions for individual patients, an error analysis was conducted to identify the factors contributing to the varying degrees of loss across the testing dataset. The analysis investigated potential correlations between loss and various factors, including autoregulation index, mean ICP values, ICP signal variance, and clustering of features in a reduced dimensionality space. This investigation aimed to identify any underlying reasons for the variation in loss across the testing dataset and to inform the development of improved models for predicting ICP in individual patients and across patients.

To investigate the relationship between the autoregulation index and loss, we computed the Pearson correlation coefficient between the ABP and ICP to obtain an autoregulation index for each prediction in the testing set for every 10 second window instance. The autoregulation index represents the degree of correspondence between ABP and ICP waveforms. We hypothesized that a higher autoregulation index would be associated with lower loss.

Next, data imbalance was assessed to examine whether poorly represented data resulted in worse predictions. This was particularly important in cases of high mean ICP, which was a small fraction of all data yet the most clinically relevant. To assess whether a high ICP was associated with poor prediction, the mean ICP value was calculated for each window of the testing dataset, and the Pearson correlation coefficient was computed between the ICP value and loss.

Additionally, to investigate the correlation between ICP signal variance and loss, the variance of the ICP signal was calculated for each instance, and the Pearson correlation coefficient was computed between the loss and the ICP signal variance.

Lastly, to examine if the quality or similarity of input features influenced loss, all three input waveforms were reduced into a two-dimensional space using the Uniform Manifold Approximation and Projection (UMAP) Python library. loss was overlaid onto these points to assess cluster-specific trends.

## V. Results

In the single-patient analysis for predicting ICP using ABP, PPG, and ECG in 10-second windows across 10 patients, the average MAE testing loss for Simple RNN was 3.17±0.68 mmHg, GRU was 1.31±0.59 mmHg, LSTM was 1.63±0.60 mmHg, TCN was 1.18±0.57 mmHg, VNET 1.22±0.57 mmHg, and Transformer was 2.65±0.82 mmHg (Table III). Model performance on for each patient can be found in Supplementary Tables 1A-1F. Based on these results, there were two tiers of model performance. GRU, LSTM, TCN, and V-NET all performed significantly better than the Simple RNN and Transformer models (refer to Supplemental Table 2A and 2B for significance testing). While there is some variation in results for GRU, LSTM, TCN, and V-NET, there is no conclusive ‘top’ model.

**Table III.** Summary of Single Patient Analysis Results

|  | Simple RNN | GRU | LSTM | TCN | V-NET | Transformer |
| --- | --- | --- | --- | --- | --- | --- |
| **Logcosh Test Loss (Train Loss) in mmHg** | $2.57 \pm 0.66$<br>($2.69 \pm 0.73$) | $0.86 \pm 0.52$<br>($0.73 \pm 0.45$) | $1.08 \pm 0.54$<br>($0.91 \pm 0.51$) | $0.75 \pm 0.50$<br>($0.41 \pm 0.26$) | $0.80 \pm 0.50$<br>($0.39 \pm 0.32$) | $2.08 \pm 0.79$<br>($2.15 \pm 0.79$) |
| **MAE Test Loss (Train Loss) in mmHg** | $3.17 \pm 0.68$<br>($3.36 \pm 0.78$) | $1.31 \pm 0.59$<br>($1.13 \pm 0.52$) | $1.63 \pm 0.60$<br>($1.38 \pm 0.62$) | $1.18 \pm 0.57$<br>($0.64 \pm 0.30$) | $1.22 \pm 0.57$<br>($0.58 \pm 0.39$) | $2.65 \pm 0.82$<br>($2.74 \pm 0.84$) |
| **RMSE Test Loss (Train Loss) in mmHg** | $4.26 \pm 0.81$<br>($4.52 \pm 0.94$) | $1.95 \pm 0.77$<br>($1.69 \pm 0.70$) | $2.26 \pm 0.89$<br>($1.87 \pm 0.84$) | $1.83 \pm 0.78$<br>($1.02 \pm 0.41$) | $1.86 \pm 0.77$<br>($0.89 \pm 0.52$) | $3.61 \pm 1.07$<br>($3.73 \pm 1.12$) |

The multi-patient analysis results show that all models perform to a similar degree with an MAE of approximately 5 mmHg (Table IV). Figure 3 shows a sample ICP waveform prediction by the TCN model. Figure 4 shows the Bland-Altman graphs for each model in the multipatient analysis with a testing set mean difference of 0.56 mmHg for Simple RNN, -0.26 mmHg for GRU, 0.09 mmHg for LSTM, 0.84 mmHg for TCN, 0.81 mmHg for V-NET, and 1.53 mmHg for Transformer. Upon further inspection of the Bland-Altman graphs for each model, it becomes clear that there is variation in model predictions. Both the Simple RNN and Transformer models have a semi-linear trend in the Bland-Altman graphs, indicating the presence of a systematic bias. In contrast, GRU, LSTM, TCN, and V-NET have no decisive correlation across the mean difference line. This trend is also observed in testing set predictions versus ground truth labels for each model (Supplementary Figure 1). There are a few lines of reasoning that may help to explain the discrepancies in model performance for the superiority of LSTM, GRU, TCN, and V-NET over the Simple RNN and Transformer. The first is the data representation itself. Transformers are particularly effective when there is a clear spatial structure in the data, such as in image or language processing tasks. They excel at capturing global dependencies but may struggle with capturing local and temporal dependencies, which are prevalent in waveform data. Similarly, Simple RNNs might have difficulty capturing long-term dependencies due to the vanishing gradient problem. On the other hand, TCN, GRU, LSTM, and V-NET architectures are designed to address these issues, making them more suitable for the task at hand with the given structure of the data. In terms of model capacity, TCN, GRU, LSTM, and V-NET architectures generally have larger model capacities compared to Simple RNN and Transformer architectures in this case. The larger capacity allows them to learn more intricate patterns and relationships in the data, which may be necessary for accurately inferring the ICP waveform. Therefore, having such discrepancies across models may elucidate insights into the nature of the computations necessary for accurate inferences.

**Table IV.**
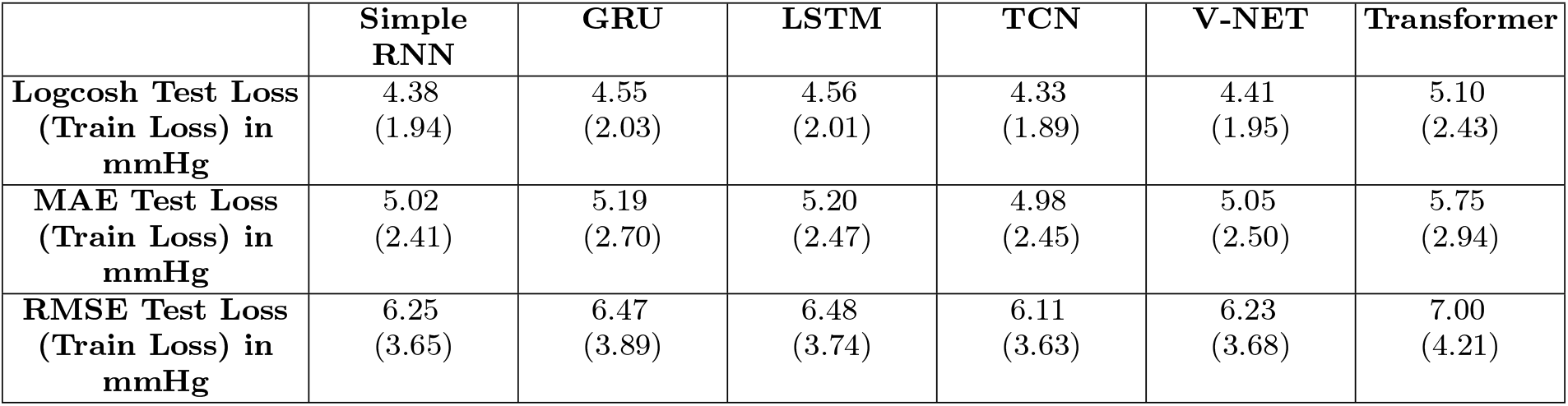
Multi-Patient Analysis Results

|  | Simple RNN | GRU | LSTM | TCN | V-NET | Transformer |
| --- | --- | --- | --- | --- | --- | --- |
| **Logcosh Test Loss (Train Loss) in mmHg** | 4.38<br>(1.94) | 4.55<br>(2.03) | 4.56<br>(2.01) | 4.33<br>(1.89) | 4.41<br>(1.95) | 5.10<br>(2.43) |
| **MAE Test Loss (Train Loss) in mmHg** | 5.02<br>(2.41) | 5.19<br>(2.70) | 5.20<br>(2.47) | 4.98<br>(2.45) | 5.05<br>(2.50) | 5.75<br>(2.94) |
| **RMSE Test Loss (Train Loss) in mmHg** | 6.25<br>(3.65) | 6.47<br>(3.89) | 6.48<br>(3.74) | 6.11<br>(3.63) | 6.23<br>(3.68) | 7.00<br>(4.21) |

**FIG. 3:**
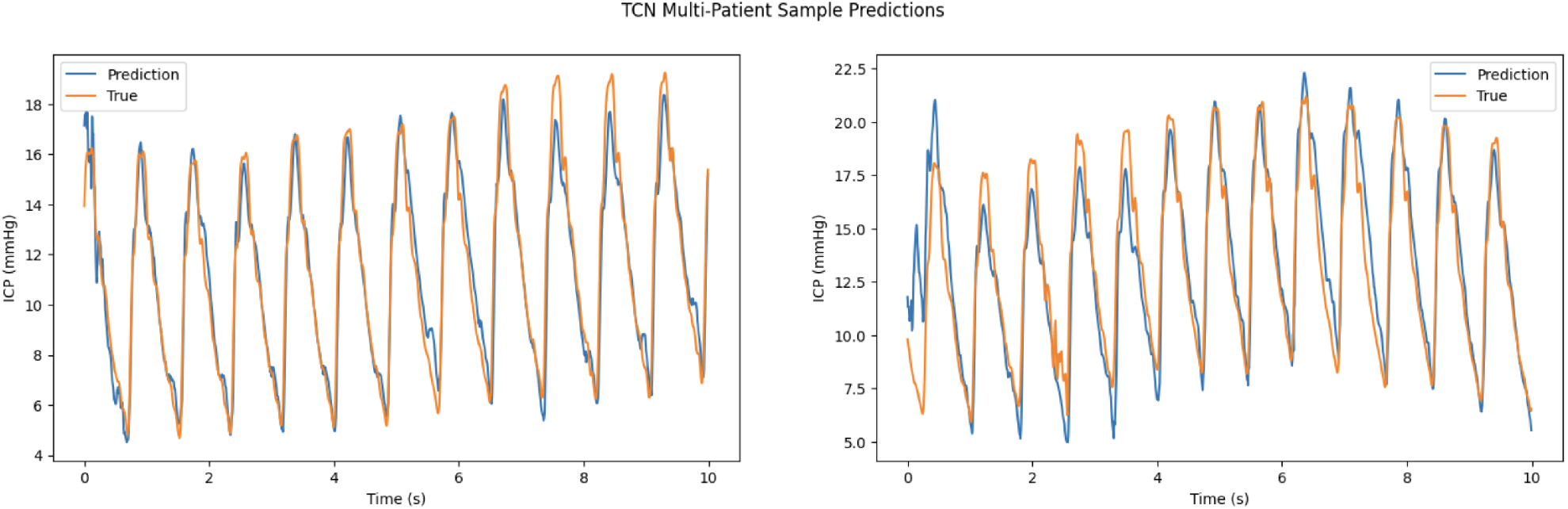
TCN Multi-Patient Waveform Prediction Visualizations

**FIG. 4:**
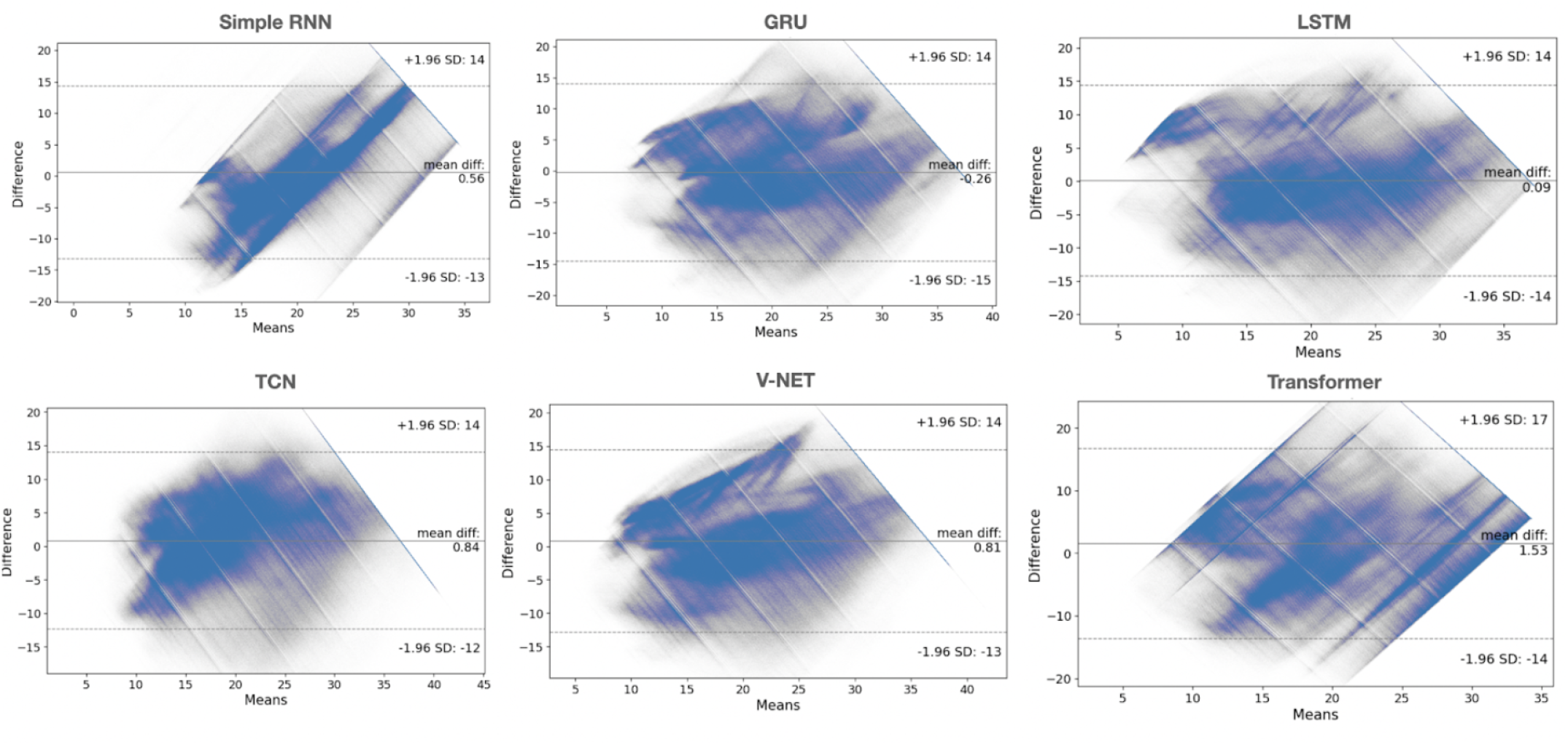
Bland-Altman Graphs for Multi-Patient Analysis for All Six Models

The ablation study for multi-patient analysis demonstrated that all three waveforms are necessary for producing the most optimal ICP prediction (Table V). When only inputting one waveform, using ECG yielded the best results across almost all models followed by PPG and ABP. When using a pair of extracranial waveforms, there was no clear trend in an optimal combination. The outlier model in the ablation study was V-NET, which showed only a small improvement in predictions with the addition of multiple extracranial waveforms. This exception to our expectations may again offer valuable insight into model architectures that are most conducive to performing accurate inferences. The V-NET architecture incorporates mechanisms and layers that enable dimensionality reduction, allowing it to capture essential information effectively. Utilizing pooling layers, the V-NET reduces spatial dimensions by downsampling feature maps. Additionally, the Conv1D layers perform 1D convolutions, capturing local patterns and dependencies, effectively reducing dimensionality and extracting relevant features, including important temporal dependencies.

**Table V.** Ablation Results for TCN Multi-Patient Analysis

| Ablation Test | RNN Test Set<br>MAE Loss<br>(mmHg) | GRU Test Set<br>MAE Loss<br>(mmHg) | LSTM Test Set<br>MAE Loss<br>(mmHg) | TCN Test Set<br>MAE Loss<br>(mmHg) | VNET Test Set<br>MAE Loss<br>(mmHg) | Transformer<br>Test Set MAE<br>Loss (mmHg) |
| --- | --- | --- | --- | --- | --- | --- |
| **ABP** | 8.23 | 7.51 | 8.14 | 8.11 | 5.42 | 8.46 |
| **PPG** | 6.83 | 7.04 | 7.20 | 7.13 | 5.21 | 8.34 |
| **ECG** | 6.57 | 6.79 | 6.72 | 6.54 | 5.24 | 8.31 |
| **ABP + PPG** | 5.23 | 6.32 | 5.78 | 5.45 | 5.17 | 7.44 |
| **ABP + ECG** | 5.91 | 5.78 | 5.66 | 5.36 | 5.32 | 6.33 |
| **PPG + ECG** | 5.12 | 5.24 | 6.18 | 6.03 | 5.06 | 6.95 |
| **ABP + PPG**<br>**+ ECG** | 5.02 | 5.19 | 5.20 | 4.98 | 5.05 | 5.75 |

Finally, in conducting our clinical error analysis, we found that model performance was not significantly correlated with autoregulation index, mean ICP values, ICP signal variance, or clustering of features in a reduced dimensionality space. Ultimately, further investigation is required to determine the causal factor for poor loss in our algorithms.

## VI. Discussion

### A. Key Results

Using extracranial continuous physiological waveform data, we present a non-invasive modeling approach for computing a surrogate ICP from extracranial waveforms across six different deep learning models. Our results suggest that deep learning algorithms can model ICP from extracranial waveforms as demonstrated by our single-patient analysis where top models (GRU, LSTM, TCN, V-NET) predicted ICP with a MAE of approximately 1.50 mmHg. Furthermore our results show our algorithms are proficient in generalizing across patients in a small training set with our top models predicting ICP with a MAE of approximately 5 mmHg with limited systematic bias. Beyond this, we demonstrate that PPG is an important waveform in predicting ICP, a novel insight. The strength of these results can be partly explained by the relationship between these extracranial waveforms and intracranial hemodynamics. In addition, these results show there exists valuable information encoded in the waveform morphologies of ECG, PPG, and ABP that relate to ICP.

There is only one study to our knowledge that aims at deriving ICP non-invasively from waveform data using transcranial doppler (TCD) [45]. This study used ABP, ECG, and cerebral blood flow velocity (derived from TCD) as input waveforms for domain adversarial neural networks and transformers to infer ICP across 11 patients with a median MAE of 3.88+/-3.26 mmHg and 3.94+/-1.71 mmHg, respectively. A limitation with this study is that it uses cerebral blood flow velocity, which is not a routinely measured waveform in the ICU given that its computation requires a TCD ultrasonography scan. As such, this data study only uses around 500,000 data points in comparison to our study which uses more than 270,000,000 data points.

We can also compare our algorithms’ performances to other on-the-market technologies that aim to measure ICP both less-invasively and invasively. TCD ultrasonography has an error range of ± 12 mmHg [46] and cannot monitor ICP continuously. Tympanic membrane displacement has an error range of ± 15 mmHg and requires a probe to be inserted into the canal of the ear [8]. Optic sheath diameter, a retinal imaging-based technique, reports a margin of error from 5-10 mmHg and also cannot monitor ICP continuously [8]. Furthermore, our top algorithms performed similarly to other state-of-the-art alternatives to the EVD such as intracranial transducers and intraparenchymal monitoring which both report a mean error of ± 7 mmHg [8,46]. While both techniques offer real-time and continuous monitoring, they are on the same level of invasiveness as an EVD, requiring surgical implantation.

### B. Novelty

This research provides compelling evidence that it is possible to accurately and in real-time compute a surrogate ICP from extracranial waveforms. With further validation, the real-time and non-invasive nature of our algorithms suggest they could be integrated into the current clinical workflow of ICP monitoring, a profound advancement towards eliminating the need for an EVD. In addition, by assessing multiple deep learning algorithms for accomplishing this task, we have also conducted a robust methodological study on the performance of deep learning algorithms on time-series data. Understanding the differences in predictions from these six deep learning algorithms could provide insights into understanding both the clinical and technical components of this multifaceted problem. Ultimately, the results of this study demonstrate that algorithmic prediction of ICP performs better than current, less invasive alternatives and on par with alternative invasive options to the EVD.

### C. Limitations

We acknowledge several limitations of this research. First, the sample size of this study was small, consisting of only 10 patients, which may impede our ability to generalize model predictions to a broader population. Second, our application of a selective pre-processing scheme to discern high-quality waveform data could introduce subjectivity and bias into the training data, potentially compromising model performance and robustness. Finally, our study lacked external validation through the use of an independent dataset, thereby restricting our ability to evaluate the generalizability and reliability of the findings beyond the data sourced from the MIMIC III Waveform Database.

### D. Future Direction

Forthcoming endeavors will concentrate on augmenting both the sample size of evaluated patients, and further examining differences in model performances. Although 10 patients were used in this study, their data quality proved invaluable in drawing clinically significant conclusions and correlations. Nevertheless, we plan to expand our inclusion criteria by accommodating missing values (presently, five-minute intervals that comprise any missing values) and adding additional patients, thereby enhancing patient representation and improving model generalizability. We will also test the models on an external dataset outside of the MIMIC III Waveform Database to assess the generalizability of model performance. In particular, we intend to externally validate our data on a more homogenous group of TBI patients using the CENTER-TBI database. Finally, we are continuing to fine tune our individual deep learning models and are examining methods for combining the strengths of each model into a ‘super-ensemble’ learner to produce more optimal results. In doing so, we seek to utilize the strengths of each model to improve patient generalizability.

### E. Supplementary Section

Supplementary Tables 1A-1F display the performance of each of the six deep learning models in all ten individual patients within the single-patient analysis. Supplementary Table 2A and 2B contains complete significance testing information when comparing these models against. Finally, Supplementary Figure 1 displays predicted versus ground truth predictions for each model in the multi-patient analysis. All tables and figures can be found after the references.

## Data Availability

All input data in this study is available online at: https://physionet.org/content/mimic3wdb/1.0/

https://physionet.org/content/mimiciii/1.4/

---

**Supplemental Table 1A.** Single Patient Analysis Results for Simple RNN

| | Pat 1 | Pat 2 | Pat 3 | Pat 4 | Pat 5 | Pat 6 | Pat 7 | Pat 8 | Pat 9 | Pat 10 | Mean ± Std | Median |
| --- | --- | --- | --- | --- | --- | --- | --- | --- | --- | --- | --- | --- |
| **Logcosh Test Loss (Train Loss) in mmHg** | 3.47<br>(3.58) | 2.49<br>(2.47) | 1.87<br>(1.85) | 2.28<br>(2.37) | 2.52<br>(2.56) | 2.36<br>(2.61) | 3.24<br>(3.29) | 3.05<br>(3.09) | 1.35<br>(1.40) | 3.11<br>(3.63) | 2.57 ± 0.66<br>(2.69 ± 0.73) | 2.51<br>(2.59) |
| **MAE Test Loss (Train Loss) in mmHg** | 4.10<br>(4.25) | 3.09<br>(3.18) | 2.46<br>(2.53) | 2.87<br>(2.94) | 3.11<br>(3.16) | 2.94<br>(3.25) | 3.86<br>(4.04) | 3.67<br>(3.83) | 1.89<br>(1.96) | 3.74<br>(4.41) | 3.17 ± 0.68<br>(3.36 ± 0.78) | 3.10<br>(3.22) |
| **RMSE Test Loss (Train Loss) in mmHg** | 5.17<br>(5.28) | 4.08<br>(4.29) | 3.07<br>(3.16) | 4.38<br>(4.64) | 4.27<br>(4.35) | 4.16<br>(4.60) | 4.92<br>(5.16) | 4.74<br>(4.96) | 2.74<br>(2.85) | 5.04<br>(5.94) | 4.26 ± 0.81<br>(4.52 ± 0.94) | 4.33<br>(4.62) |

**Supplemental Table 1B.** Single Patient Analysis Results for GRU

| | Pat 1 | Pat 2 | Pat 3 | Pat 4 | Pat 5 | Pat 6 | Pat 7 | Pat 8 | Pat 9 | Pat 10 | Mean ± Std | Median |
| --- | --- | --- | --- | --- | --- | --- | --- | --- | --- | --- | --- | --- |
| **Logcosh Test Loss (Train Loss) in mmHg** | 2.05<br>(1.81) | 0.92<br>(0.83) | 0.32<br>(0.24) | 1.06<br>(0.87) | 0.83<br>(0.53) | 0.86<br>(0.89) | 0.53<br>(0.57) | 0.40<br>(0.34) | 0.40<br>(0.38) | 1.24<br>(0.84) | 0.86 ± 0.52<br>(0.73 ± 0.45) | 0.85<br>(0.70) |
| **MAE Test Loss (Train Loss) in mmHg** | 2.63<br>(2.32) | 1.42<br>(1.28) | 0.67<br>(0.47) | 1.54<br>(1.40) | 1.32<br>(0.83) | 1.30<br>(1.39) | 0.94<br>(0.96) | 0.78<br>(0.73) | 0.79<br>(0.72) | 1.75<br>(1.17) | 1.31 ± 0.59<br>(1.13 ± 0.52) | 1.31<br>(1.07) |
| **RMSE Test Loss (Train Loss) in mmHg** | 3.48<br>(3.10) | 2.03<br>(1.84) | 1.07<br>(0.80) | 2.59<br>(2.37) | 1.89<br>(1.19) | 2.03<br>(2.16) | 1.32<br>(1.44) | 1.26<br>(1.13) | 1.23<br>(1.11) | 2.59<br>(1.74) | 1.95 ± 0.77<br>(1.69 ± 0.70) | 1.96<br>(1.59) |

**Supplemental Table 1C.** Single Patient Analysis Results for LSTM

| | Pat 1 | Pat 2 | Pat 3 | Pat 4 | Pat 5 | Pat 6 | Pat 7 | Pat 8 | Pat 9 | Pat 10 | Mean ± Std | Median |
| --- | --- | --- | --- | --- | --- | --- | --- | --- | --- | --- | --- | --- |
| **Logcosh Test Loss (Train Loss) in mmHg** | 2.02<br>(1.74) | 0.90<br>(0.54) | 0.48<br>(0.43) | 1.00<br>(0.91) | 0.83<br>(0.52) | 0.89<br>(0.75) | 0.47<br>(0.39) | 0.83<br>(0.78) | 1.62<br>(1.33) | 1.80<br>(1.73) | 1.08 ± 0.54<br>(0.91 ± 0.51) | 0.90<br>(0.77) |
| **MAE Test Loss (Train Loss) in mmHg** | 2.61<br>(2.27) | 1.38<br>(0.82) | 0.88<br>(0.81) | 1.47<br>(1.41) | 1.32<br>(0.81) | 1.34<br>(1.17) | 0.88<br>(0.67) | 1.91<br>(1.71) | 2.15<br>(1.79) | 2.36<br>(2.29) | 1.63 ± 0.60<br>(1.38 ± 0.62) | 2.04<br>(1.50) |
| **RMSE Test Loss (Train Loss) in mmHg** | 3.45<br>(2.98) | 2.02<br>(1.21) | 1.29<br>(1.27) | 2.57<br>(2.47) | 1.90<br>(1.16) | 2.05<br>(1.73) | 1.24<br>(0.97) | 1.31<br>(1.21) | 3.34<br>(2.43) | 3.39<br>(3.23) | 2.26 ± 0.89<br>(1.87 ± 0.84) | 2.04<br>(1.50) |

**Supplemental Table 1D.**
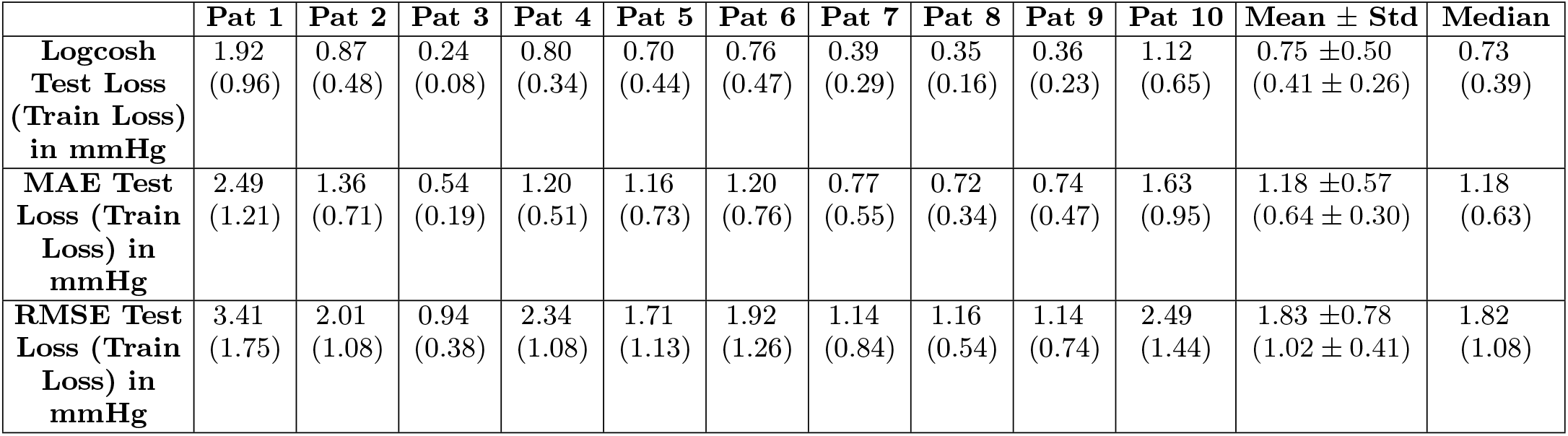
Single Patient Analysis Results for TCN

| | Pat 1 | Pat 2 | Pat 3 | Pat 4 | Pat 5 | Pat 6 | Pat 7 | Pat 8 | Pat 9 | Pat 10 | Mean ± Std | Median |
| --- | --- | --- | --- | --- | --- | --- | --- | --- | --- | --- | --- | --- |
| **Logcosh Test Loss (Train Loss) in mmHg** | 1.92<br>(0.96) | 0.87<br>(0.48) | 0.24<br>(0.08) | 0.80<br>(0.34) | 0.70<br>(0.44) | 0.76<br>(0.47) | 0.39<br>(0.29) | 0.35<br>(0.16) | 0.36<br>(0.23) | 1.12<br>(0.65) | 0.75 ± 0.50<br>(0.41 ± 0.26) | 0.73<br>(0.39) |
| **MAE Test Loss (Train Loss) in mmHg** | 2.49<br>(1.21) | 1.36<br>(0.71) | 0.54<br>(0.19) | 1.20<br>(0.51) | 1.16<br>(0.73) | 1.20<br>(0.76) | 0.77<br>(0.55) | 0.72<br>(0.34) | 0.74<br>(0.47) | 1.63<br>(0.95) | 1.18 ± 0.57<br>(0.64 ± 0.30) | 1.18<br>(0.63) |
| **RMSE Test Loss (Train Loss) in mmHg** | 3.41<br>(1.75) | 2.01<br>(1.08) | 0.94<br>(0.38) | 2.34<br>(1.08) | 1.71<br>(1.13) | 1.92<br>(1.26) | 1.14<br>(0.84) | 1.16<br>(0.54) | 1.14<br>(0.74) | 2.49<br>(1.44) | 1.83 ± 0.78<br>(1.02 ± 0.41) | 1.82<br>(1.08) |

**Supplemental Table 1E.** Single Patient Analysis Results for V-NET

| | Pat 1 | Pat 2 | Pat 3 | Pat 4 | Pat 5 | Pat 6 | Pat 7 | Pat 8 | Pat 9 | Pat 10 | Mean ± Std | Median |
| --- | --- | --- | --- | --- | --- | --- | --- | --- | --- | --- | --- | --- |
| **Logcosh Test Loss (Train Loss) in mmHg** | 1.94<br>(1.16) | 0.88<br>(0.48) | 0.25<br>(0.05) | 0.90<br>(0.47) | 0.75<br>(0.43) | 0.75<br>(0.34) | 0.41<br>(0.17) | 0.37<br>(0.13) | 0.37<br>(0.16) | 1.17<br>(0.48) | 0.80 ± 0.50<br>(0.39 ± 0.32) | 0.75<br>(0.39) |
| **MAE Test Loss (Train Loss) in mmHg** | 2.52<br>(1.49) | 1.36<br>(0.69) | 0.57<br>(0.11) | 1.34<br>(0.75) | 1.23<br>(0.68) | 1.18<br>(0.53) | 0.80<br>(0.28) | 0.75<br>(0.26) | 0.76<br>(0.37) | 1.68<br>(0.67) | 1.22 ± 0.57<br>(0.58 ± 0.39) | 1.21<br>(0.61) |
| **RMSE Test Loss (Train Loss) in mmHg** | 3.38<br>(1.94) | 2.00<br>(1.05) | 0.98<br>(0.19) | 2.45<br>(1.37) | 1.78<br>(0.95) | 1.90<br>(0.86) | 1.19<br>(0.42) | 1.21<br>(0.44) | 1.15<br>(0.58) | 2.54<br>(1.07) | 1.86 ± 0.77<br>(0.89 ± 0.52) | 1.84<br>(0.91) |

**Supplemental Table 1F.** Single Patient Analysis Results for Transformer

| | Pat 1 | Pat 2 | Pat 3 | Pat 4 | Pat 5 | Pat 6 | Pat 7 | Pat 8 | Pat 9 | Pat 10 | Mean ± Std | Median |
| --- | --- | --- | --- | --- | --- | --- | --- | --- | --- | --- | --- | --- |
| **Logcosh Test Loss (Train Loss) in mmHg** | 3.21<br>(3.30) | 1.91<br>(2.03) | 0.92<br>(0.94) | 2.41<br>(2.56) | 1.57<br>(1.66) | 3.21<br>(3.25) | 2.07<br>(2.11) | 1.65<br>(1.83) | 1.19<br>(1.19) | 2.63<br>(2.62) | 2.08 ± 0.79<br>(2.15 ± 0.79) | 1.99<br>(2.07) |
| **MAE Test Loss (Train Loss) in mmHg** | 3.84<br>(3.95) | 2.50<br>(2.66) | 1.43<br>(1.45) | 2.99<br>(3.23) | 2.13<br>(2.24) | 3.82<br>(3.87) | 2.65<br>(2.71) | 2.21<br>(2.41) | 1.71<br>(1.71) | 3.22<br>(3.21) | 2.65 ± 0.82<br>(2.74 ± 0.84) | 2.58<br>(2.69) |
| **RMSE Test Loss (Train Loss) in mmHg** | 4.87<br>(5.01) | 3.34<br>(3.56) | 1.90<br>(1.83) | 4.53<br>(4.89) | 2.91<br>(3.08) | 5.00<br>(5.07) | 3.58<br>(3.66) | 3.00<br>(3.27) | 2.46<br>(2.46) | 4.51<br>(4.50) | 3.61 ± 1.07<br>(3.73 ± 1.12) | 3.46<br>(3.61) |

**Supplemental Table 2A.**
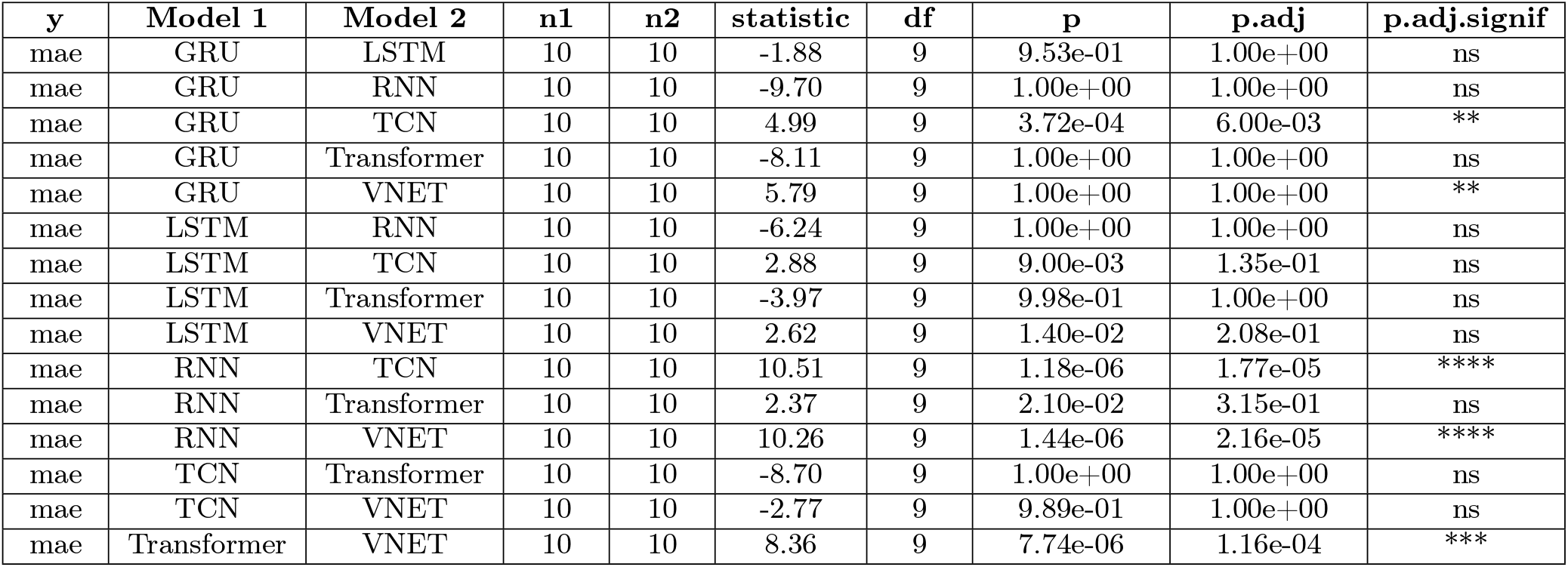
Single Patient Paired One-Tailed T-Test Between Models (Model 1 MAE > Model 2 MAE)

**Supplemental Table 2B.** Single Patient Paired One-Tailed T-Test Between Models (Model 1 MAE < Model 2 MAE)

| y | Model 1 | Model 2 | n1 | n2 | statistic | df | p | p.adj | p.adj.signif |
| --- | --- | --- | --- | --- | --- | --- | --- | --- | --- |
| mae | GRU | LSTM | 10 | 10 | -1.88 | 9 | 4.70e-02 | 7.02e-01 | ns |
| mae | GRU | RNN | 10 | 10 | -9.70 | 9 | 2.31e-06 | 3.46e-05 | **** |
| mae | GRU | TCN | 10 | 10 | 4.99 | 9 | 1.00e+00 | 1.00e+00 | ns |
| mae | GRU | Transformer | 10 | 10 | -8.11 | 9 | 9.96e-06 | 1.49e-04 | *** |
| mae | GRU | VNET | 10 | 10 | 5.79 | 9 | 1.00e+00 | 1.00e+00 | ns |
| mae | LSTM | RNN | 10 | 10 | -6.24 | 9 | 7.59e-05 | 1.00e-03 | ** |
| mae | LSTM | TCN | 10 | 10 | 2.88 | 9 | 9.91e-01 | 1.00e+00 | ns |
| mae | LSTM | Transformer | 10 | 10 | -3.97 | 9 | 2.00e-03 | 2.40e-02 | * |
| mae | LSTM | VNET | 10 | 10 | 2.62 | 9 | 9.86e-01 | 1.00e+00 | ns |
| mae | RNN | TCN | 10 | 10 | 10.51 | 9 | 1.00e+00 | 1.00e+00 | ns |
| mae | RNN | Transformer | 10 | 10 | 2.37 | 9 | 9.79e-01 | 1.00e+00 | ns |
| mae | RNN | VNET | 10 | 10 | 10.26 | 9 | 1.00e+00 | 1.00e+00 | ns |
| mae | TCN | Transformer | 10 | 10 | -8.70 | 9 | 5.64e-06 | 8.46e-05 | **** |
| mae | TCN | VNET | 10 | 10 | -2.77 | 9 | 1.10e-02 | 1.64e-01 | ns |
| mae | Transformer | VNET | 10 | 10 | 8.36 | 9 | 1.00e+00 | 1.00e+00 | ns |

**Supplemental Figure 1.**
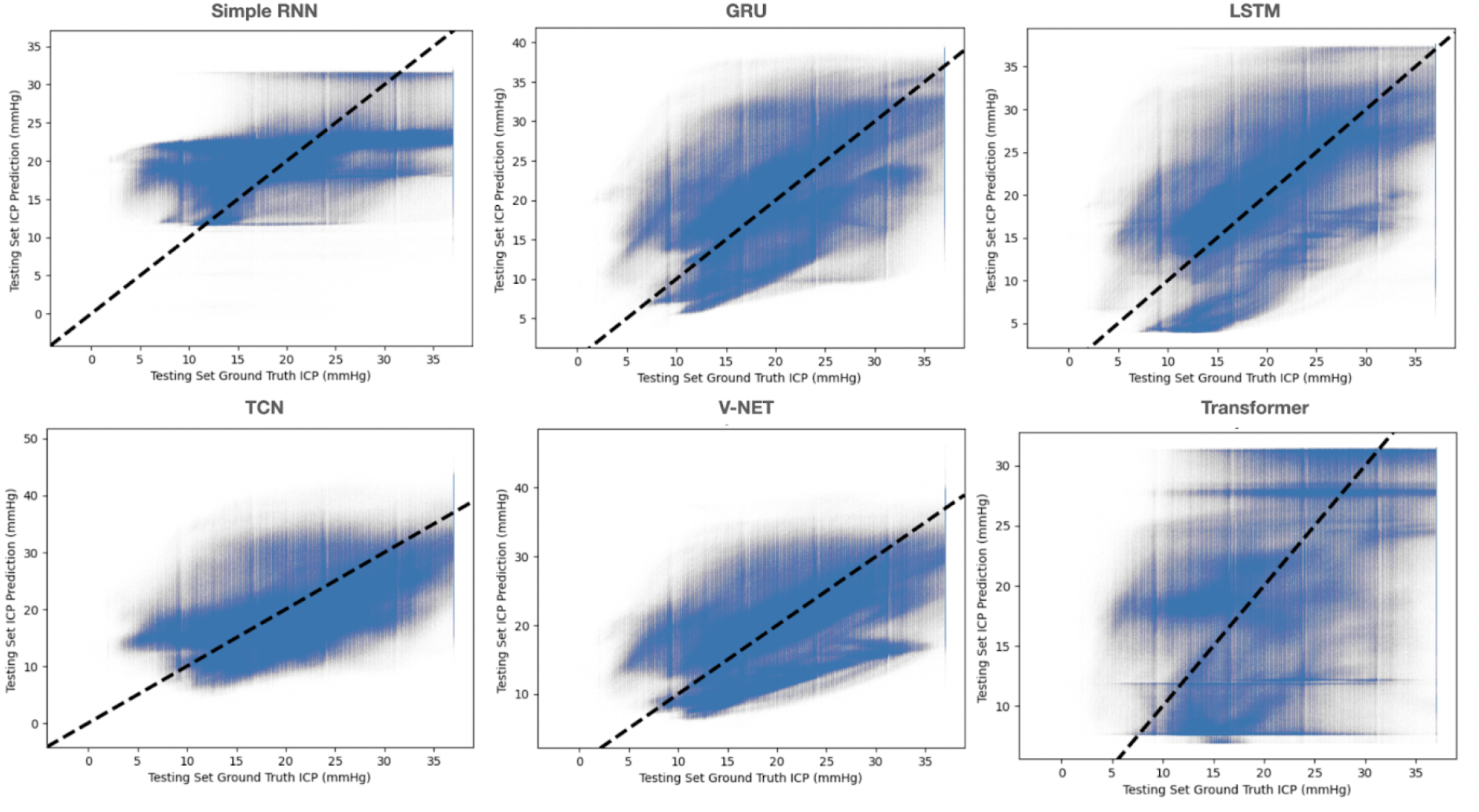
Multi-Train Analysis Prediction vs Ground Truth for Each Model

